# Evaluation of SARS-CoV-2 neutralization assays for antibody monitoring in natural infection and vaccine trials

**DOI:** 10.1101/2020.12.07.20245431

**Authors:** Anton M. Sholukh, Andrew Fiore-Gartland, Emily S. Ford, Yixuan J. Hou, Victor Tse, Hannah Kaiser, Arnold Park, Florian A. Lempp, Russell Saint Germain, Emily Bossard, Jia Jin Kee, Kurt Diem, Andrew B. Stuart, Peter B. Rupert, Chance Brock, Matthew Buerger, Margaret K. Doll, April Kaur Randhawa, Leonidas Stamatatos, Roland K. Strong, Colleen McLaughlin, Keith R. Jerome, Ralph S. Baric, David Montefiori, Lawrence Corey

## Abstract

Determinants of protective immunity against SARS-CoV-2 infection require the development of well-standardized, reproducible antibody assays to be utilized in concert with clinical trials to establish correlates of risk and protection. This need has led to the appearance of a variety of neutralization assays used by different laboratories and companies. Using plasma samples from COVID-19 convalescent individuals with mild-to-moderate disease from a localized outbreak in a single region of the western US, we compared three platforms for SARS-CoV-2 neutralization: assay with live SARS-CoV-2, pseudovirus assay utilizing lentiviral (LV) and vesicular stomatitis virus (VSV) packaging, and a surrogate ELISA test. Vero, Vero E6, HEK293T cells expressing human angiotensin converting enzyme 2 (hACE2), and TZM-bl cells expressing hACE2 and transmembrane serine protease 2 (TMPRSS2) were evaluated. Live-virus and LV-pseudovirus assay with HEK293T cells showed similar geometric mean titers (GMTs) ranging 141–178, but VSV-pseudovirus assay yielded significantly higher GMT (310 95%CI 211-454; p < 0.001). Fifty percent neutralizing dilution (ND50) titers from live-virus and all pseudovirus assay readouts were highly correlated (Pearson *r* = 0.81–0.89). ND50 titers positively correlated with plasma concentration of IgG against SARS-CoV-2 spike and receptor binding domain (RBD) (*r* = 0.63–0.89), but moderately correlated with nucleoprotein IgG (*r* = 0.46–0.73). There was a moderate positive correlation between age and spike (Spearman’s rho=0.37, p=0.02), RBD (rho=0.39, p=0.013) and nucleoprotein IgG (rho=0.45, p=0.003). ND80 showed stronger correlation with age than ND50 (ND80 rho=0.51 (p=0.001), ND50 rho=0.28 (p=0.075)). Our data demonstrate high concordance between cell-based assays with live and pseudotyped virions.

## INTRODUCTION

The coronavirus disease 2019 (COVID-19) pandemic, caused by severe acute respiratory syndrome coronavirus 2 (SARS-CoV-2), has caused more than 63 million confirmed infections and over 1.4 million deaths worldwide as of November 30^th^, 2020 (https://www.worldometers.info/coronavirus). To minimize virus transmission and reduce mortality, regulations enforcing mask use and limiting close contact were implemented in many countries. An initial success of these orders with an observable reduction in cases led to relaxed social distancing orders and lockdowns. This in turn has been followed by a new surge of infections, with case rates and mortality exceeding the initial outbreaks. It is clear that the reopening of society and return to a pre-pandemic lifestyle will be possible only after safe and efficacious vaccines and therapies are developed and implemented.

The efficacy of most licensed vaccines correlates with pathogen-neutralizing antibodies elicited by vaccination (1). Humans can mount neutralizing antibody (nAb) responses against SARS-CoV-2 during natural infection (2–5). Epidemiologic data suggest that reinfection rates seem low, albeit increasing numbers of sporadic reinfections are being reported (6, 7). A crucial unknown at this time is what immune responses are associated with protective immunity. Recent studies suggest passive infusion of monoclonal antibodies can alter COVID-19 disease progression. Infusion of convalescent sera is more controversial regarding its efficacy. In order to determine what constitutes protective immunity in human populations, well-standardized, reproducible antibody assays are required to establish correlates of risk and protection. The current study evaluates several assays that are under validation for use to determine correlates of protection in vaccine studies evaluating immune responses in persons with symptomatic COVID-19. These assays include a live-virus neutralization assay, which is notable because work with live SARS-CoV-2 requires Biosafety Level 3 (BSL-3) containment; something not readily available at many institutions and not easily amenable to high-throughput experiments.

To avoid these barriers and improve assay feasibility, a variety of live-virus neutralization assays use recombinant SARS-CoV-2 (rSARS-CoV-2) containing GFP or luciferase reporter genes at the *ORF7* locus of the viral genome (8, 9). These recombinant viruses replicate similarly to SARS-CoV-2 clinical isolates in vitro and successfully infect primary airway epithelial cell cultures. A fluorescence-based rSARS-CoV-2 neutralization assay yielded comparable results to plaque reduction neutralization test (PRNT) in nAb detection from convalescent patient plasma (8). With a shorter turnaround time (24-48 hours for reporter virus vs. 3 days for PRNT), rSARS-CoV-2 provides a useful high-throughput platform to study nAb responses, but still requires a BSL-3 laboratory for assay set up and readout.

Reporter assays using pseudotyped viruses, which are restricted to a single round of intracellular replication, allow experiments to be carried out in BSL-2 environments. Pseudotyped viral particles can be created with two packaging platforms: lentiviral/retroviral and vesicular stomatitis virus (VSV) (10–13). Both packaging systems have been used for SARS-CoV-2 pseudovirus reporter neutralization assays and have been tested on multiple cell types expressing the SARS-CoV-2 entry receptor angiotensin converting enzyme 2 (ACE2) (14–16). Human lung carcinoma Calu-3 cells, colorectal adenocarcinoma Caco-2 cells, and green monkey kidney epithelial Vero cells were the most susceptible for VSV-pseudovirus entry (17). Both Calu-3 and Caco-2, but not Vero, cells express endogenous transmembrane serine protease 2 (TMPRSS2), a protease that facilitates SARS-CoV-2 cell entry; however, there were no significant differences in VSV-pseudovirus entry between the three cell types (17). Vero E6 cells have the highest level of ACE2 expression and both Vero and Vero E6 cells exogenously expressing TMPRSS2 were highly susceptible to SARS-CoV-2 infection (17–19). HEK 293T cells transfected to express human ACE2 were also developed and tested in pseudovirus neutralization assays (20).

Binding antibody (bAb) assays are an alternative to cell-based SARS-CoV-2 neutralization assays (15, 21, 22). Major advantages of bAb assays include their cost, speed and safety. The receptor binding domain (RBD) of the SARS-CoV-2 spike glycoprotein is the major target for nAbs (23–25), and bAb assays are therefore based on the detection of antibodies that compete with ACE2 for RBD binding. As opposed to measuring actual virus neutralization, surrogate bAb assays report percent binding inhibition between RBD and ACE2, which is then interpreted as percent neutralization. Competitive surface plasmon resonance (SPR) (15) and ELISA (21, 22) have both been developed for this purpose. While they provide inexpensive and rapid detection of RBD-targeting nAbs, bAb assays cannot measure neutralization via non-RBD spike protein epitopes or concerted action of antibodies targeting different epitopes.

In the current study, we used plasma samples from convalescent individuals with mild-to-moderate COVID-19 disease to compare three different SARS-CoV-2 neutralization platforms: 1) a live recombinant SARS-CoV-2 assay, 2) pseudovirus-based assays, and 3) a surrogate, ELISA-based test. In addition, two different pseudovirus packaging systems and three different cell lines were characterized. We also examined the correlation between neutralization and the concentration of SARS-CoV-2 nucleoprotein-, spike- and RBD-specific IgG.

## METHODS

### Study Population & Specimen Collection

Residents of Blaine County, Idaho ≥18 years of age volunteered for study participation by completing a secure online intake form. Study volunteers were selected for participation using a two-phased approach. In the first phase, study volunteers were randomly selected for participation after stratification by ZIP code, and within ZIP code, age, gender and race/ethnicity. In the second phase, all volunteers who were Blaine County first responders and their families were selected for participation. Selected volunteers were invited for study participation in May 2020 and sent an electronic consent statement. Following enrollment, participants completed a questionnaire in REDCap, a secure software tool, to collect demographic information and symptom histories since January 15, 2020. Appointments for sample collection were scheduled upon survey completion. Blood was collected into 10 mL vials containing acid citrate dextrose and shipped overnight to the Fred Hutchinson Cancer Research Center. Plasma was separated by centrifugation at 1200×g for 15 mins and aliquoted into cryovials. One aliquot was submitted for Architect SARS-CoV-2 IgG assay (Abbott). Others were heat inactivated for 30 min at 56 °C, frozen at −80 °C and distributed to testing laboratories. Study participants were informed of the qualitative results of the Abbott IgG Serology assay via email within one week since obtaining test results. This study was approved by the Fred Hutch Institutional Review Board and all study materials were provided in both English and Spanish.

### Protein antigens

A recombinant form of a synthetic construct (SARS_CoV_2_ectoCSPP (26); GenBank: QJE37812.1) of the spike (S) glycoprotein from SARS-CoV-2 Wuhan-Hu-1 was produced in human HEK293 cells (FreeStyle™ 293-F Cells, ThermoFisher, Waltham, MA) using a lentivirus expression system (27) and purified by nickel affinity and size-exclusion chromatography. Purity and solution monodispersivity were confirmed by comparative reduced/non-reduced PAGE, analytical size-exclusion chromatography, and static/dynamic light scattering on Uncle (Unchained Labs, Pleasanton, CA) and showed uniform trimerization. The recombinant protein was modified by replacing the native leader sequence with a murine Igk leader, removing the polybasic S1/S2 cleavage site (RRAR to A), stabilized with a pair of proline mutations (2P), and incorporating a thrombin cleavage site, a T4 foldon trimerization domain, a hexa-histidine purification tag, and a C-terminal Avi-Tag (28). After purification, the protein was sterile filtered and aliquoted in DPBS, no calcium, no magnesium (ThermoFisher). An analogous RBD-only version of this construct, swapping a TEV protease (29) site for the Thrombin site, was also produced following identical protocols. Alternatively, SARS-CoV-2 spike glycoprotein was produced as described elsewhere (23). Both spike protein preparations were tested in binding assay and no difference in recognition by serum and plasma samples from different convalescent subjects was found. SARS-CoV-2 nucleoprotein was purchased from GenScript (Piscataway, NJ) and tetanus toxoid from Lonza (Basel, Switzerland).

### VSV-pseudovirus

The codon-optimized sequence of the SARS-CoV-2 spike protein (YP_009724390.1) with a truncation of the 19 C-terminal amino acids (D19) was cloned into a pcDNA3.1(+) vector (ThermoFisher) under control of the human CMV promoter to generate pcDNA3.1(+)-SARS-CoV-2-D19. The C-terminal truncation leads to a deletion of the ER-retention signal, localizing the spike protein to the cell surface, which enhances pseudovirus packaging (30). VSV(G*ΔG-luciferase) system was purchased from Kerafast (13, 31). Twenty-four hours prior infection with VSV(G*ΔG-luciferase), 293T cells were transfected with pcDNA-WuhanCoV-S-D19. Next day, supernatant was harvest, centrifuged for 5 min at 1,000xg, aliquoted and stored at −80 °C. TCID_50_ was measured by infecting Vero cells (catalog number CCL-81; ATCC) with serial 2-fold dilutions of the prepared pseudovirus.

### LV-pseudovirus

An expression plasmid encoding codon-optimized full-length spike of the Wuhan-1 strain (VRC7480), was provided by Drs. Barney Graham and Kizzmekia Corbett at the Vaccine Research Center, National Institutes of Health (USA). The D614G mutation was introduced into VRC7480 by site-directed mutagenesis using the QuikChange Lightning Site-Directed Mutagenesis Kit from (catalog number 210518; Agilent Technologies). The mutation was confirmed by full-length spike gene sequencing. Pseudovirions were produced in HEK 293T/17 cells (catalog number CRL-11268; ATCC) by transfection using Fugene 6 (catalog number E2692; Promega). Pseudovirions for 293T/ACE2 infection were produced by co-transfection with a lentiviral backbone (pCMV-ΔR8.2) and firefly luciferase reporter gene (pHR’-CMV-Luc) (32). Pseudovirions for TZM-bl/ACE2/TMPRSS2 infection were produced by co-transfection with the Env-deficient lentiviral backbone pSG3ΔEnv (kindly provided by Drs Beatrice Hahn and Feng Gao). Culture supernatants from transfections were clarified of cells by low-speed centrifugation and filtration (0.45 µm filter) and stored in 1 ml aliquots at −80°C.

### Detection of IgG antibodies to SARS-CoV-2 using a commercial serologic assay

Plasma samples were tested at the Clinical Laboratory Improvement Amendments (CLIA)-certified University of Washington Virology lab using the Architect SARS-CoV-2 IgG assay (Abbott) under the Food and Drug Administration’s Emergency Use Authorization. The assay is a chemiluminescent microparticle immunoassay that measures IgG antibodies to the SARS-CoV-2 nucleocapsid protein. Qualitative results and index values reported by the instrument were used in analyses. Recommended index value cutoff of 1.40 was used for determining positivity (33).

### Luminex SARS-CoV-2 IgG binding antibody assay

Protein antigens were coupled to the Bio-Plex Pro Magnetic COOH beads in a ratio of 10 μg of antigen per 2.5 x 10^6^ beads in a two-step carbodiimide reaction. First, beads were washed and resuspended in Activation Buffer (100 mM MES, pH 6) and then incubated with N-hydroxysulfosuccinimide (Sulfo-NHS, catalog number 24520; ThermoFisher) and 1-ethyl-3-[3-dimethlyaminopropyl]carbodiimide-HCl (EDC, catalog number 77149; ThermoFisher) also dissolved in Activation Buffer for 20 minutes on an end-over-end rotational mixer at room temperature protected from light. Activated beads were washed three times in Activation buffer. For coupling, antigen was mixed with activated beads and reaction was carried out for 2 h on a rotational mixer at room temperature protected from light. Conjugated beads were washed three times with Wash buffer (PBS, 0.05% Tween-20, 1% BSA, 0.1% NaN_3_) and finally resuspended in Wash buffer at 10^7^ beads/ml. Beads were stored at 4 °C for no longer than 30 days.

Antigen-specific IgG was measured using two replicate dilutions. Beads were blocked with phosphate buffered saline (PBS; Gibco) containing 5% Blotto (Bio-Rad) and 0.05% Tween-20 (Sigma) and incubated for 1 hour with serially diluted plasma samples. Next, beads were washed 3 times with 0.05% Tween-20 in PBS and incubated with anti-human IgG Fc-PE (catalog number 2048-09; Sothern Biotech). After incubation with secondary antibody, beads were washed and resuspended in PBS with 1% BSA and 0.05% Tween-20 and binding data were collected on Bio-Plex 200 instrument (Bio-Rad). Median Fluorescence Intensity (MFI) was measured for a minimum of 50 beads per region. Background was established by measuring the MFI of beads conjugated to antigens but incubated in Assay buffer. Background MFI values were subtracted from all readings. We also trialed unconjugated beads and beads conjugated to a decoy antigen with the same plasma samples used in testing and did not detect non-specific binding above the assay background described above. Pooled sera from normal human donors collected in 2015 – 2016 was included as the negative control for SARS-CoV-2 antigens. For the positive control we used convalescent plasma from a subject with PCR-confirmed severe COVID-19.

An IgG standard curve run in duplicate was used to estimate IgG concentration. Anti-human IgG Fab-specific (Southern Biotech) was conjugated to the same bead regions used to conjugate to antigen proteins. IgG-coupled beads were blocked, washed and incubated with serially diluted human standard IgG (catalog number I4506; Sigma) for 1 h. Standard beads were washed and incubated with anti-human IgG Fc-PE and MFI was measured as described above. MFI readings and associated IgG concentrations were fitted to a four-parameter logistic curve (4PL) using the R packages *nCal* and *drc*. A standard curve for each experiment was used to obtain the effective concentrations of IgG in serum using the MFI measured with antigen-coated beads. Since serum samples were also run as a dilution series we used the median of the estimated concentrations from the dilutions that yielded MFIs between 100 and 10,000. Serum with all values above (below) this range were right (left) censored at the concentration of the minimum (maximum) MFI.

### Live SARS-CoV-2 neutralization assay

Assay was carried out in BSL-3 suite. Vero E6 cells were seeded at 2×10^4^ cells/well in a 96-well plate 24 h before the assay. Seventy five pfu of the recombinant SARS-CoV-2-nanoLuc virus (rSARS-CoV-2-nLuc) (9) were mixed with Ab at 1:1 ratio and incubated at 37°C for 1h. A 8-points, 3-fold dilution curve was generated for each sample with starting concentration at 1:50. Virus and Ab mix was added to each well and incubated at 37°C + 5% CO_2_ for 48h. Luciferase activities were measured by Nano-Glo Luciferase Assay System (Promega) following manufacturer protocol using SpectraMax M3 luminometer (Molecular Devices). Percent neutralization was calculated by the following equation: [1-(RLU with sample/ RLU with mock treatment)] x 100%.

### VSV pseudovirus neutralization assay

Vero cells (ATCC® CCL-81™) were seeded at 2×10^4^ cells/well in a black-walled 96-well plates 24 hours before the assay. A 7-point, 3-fold dilution curve was generated with starting sample dilution at 1:20 in a separate round-bottom 96-well plate. 3.8×10^2^ TCID50 of rVSV(G*ΔG-luciferase) pseudovirus with SARS-CoV-2-D19 spike protein (PsVSV-Luc-D19) was mixed with the plasma dilutions. Plasma-virus mixture was incubated at 37 °C in 5% CO_2_ for 30 minutes. After incubation, plasma-virus mixture was transferred onto the Vero cells. Cells were then incubated at 37 °C, 5% CO_2_ for 18-20 hours. Luciferase activity was measured by Bio-Glo Luciferase Assay System (catalog number G7940; Promega) following manufacturer protocol using 2030 VICTOR X3 multilabel reader (PerkinElmer). Percent virus neutralization was calculated by the following equation: [1-(luminescence of sample/ luminescence of cells+virus control)] x 100%. All the live virus experiments were performed under BSL-3 conditions at negative pressure, by operators in Tyvek suits wearing personal powered-air purifying respirators.

### LV-pseudovirus neutralization assays

Neutralization of SARS-CoV-2 Spike-pseudotyped virus was performed by using lentiviral vectors and infection in either HEK 293T cells expressing human ACE2 (293T/ACE2.MF) or TZM-bl cells expressing both ACE2 and TMPRSS2 (TZM-bl/ACE2/TMPRSS2 cells). Both cell lines kindly provided by Drs. Mike Farzan and Huihui Mu at Scripps). Cells were maintained in DMEM containing 10% FBS, 1% Pen Strep and 3 ug/ml puromycin.

### 293T/ACE2 cells pseudovirus assay

For the 293T/ACE2 assay, a pre-titrated dose of virus was incubated with serial 3-fold dilutions of test sample in duplicate in a total volume of 150 ul for 1 hr at 37°C in 96-well flat-bottom black/white culture plates. Freshly trypsinized cells (10,000 cells in 100 ul of growth medium) was added to each well. One set of control wells received cells + virus (virus control) and another set received cells only (background control). After 68-72 hours of incubation, 100 ul of cell lysate was transferred to a 96-well black/white plate (catalog number 6005060; Perkin-Elmer) for measurements of luminescence using the Promega Luciferase Assay System (catalog number E1501; Promega). Neutralization titers are the serum dilution at which RLUs were reduced by 50% and 80% compared to virus control wells after subtraction of background RLUs. MPI is the reduction in RLU at the lowest serum dilution tested.

### ACE2/TMPRSS2 TZM-bl cells pseudovirus assay

For the TZM-bl/ACE2/TMPRSS2 assay, a pre-titrated dose of virus was incubated with serial 3-fold dilutions of test sample in duplicate in a total volume of 150 ul for 1 hr at 37°C in 96-well flat-bottom culture plates. Freshly trypsinized cells (10,000 cells in 100 ul of growth medium containing 75 ug/ml DEAE dextran) were added to each well. One set of control wells received cells + virus (virus control) and another set received cells only (background control). After 68-72 hours of incubation, 100 ul of cell lysate was transferred to a 96-well black solid plate (Costar) for measurements of luminescence using the BriteLite Luminescence Reporter Gene Assay System (PerkinElmer Life Sciences). Neutralization titers are the serum dilution at which relative luminescence units (RLU) were reduced by 50% and 80% compared to virus control wells after subtraction of background RLUs. Maximum percent inhibition (MPI) is the reduction in RLU at the lowest serum dilution tested.

### SARS-CoV-2 Surrogate Virus Neutralization Test (sVNT)

Assay was performed according to manufacturer (GenScript) protocol and recommendations as follows. Capture plate was incubated with plasma samples diluted 1:10, washed and probed with secondary antibody. Assay was developed via TMB (ThermoFisher) and OD at 450 nm was measured using SpectraMax M2 reader (Molecular Devices). Positive and negative controls were provided in the kit. Binding inhibition was determined via the following formula: Inhibition = (1 – (OD of sample / OD of Negative control)) × 100%. Percent binding inhibition was interpreted as a percent neutralization. In order to determine ND50, plasma samples were serially diluted starting from 1:10 and assay was performed as described above.

### Statistical Analysis and Visualization

Neutralization titers were defined as the plasma dilution that reduced relative luminescence units (RLU) by 50% or 80% relative to virus control wells (cells + virus only) after subtraction of background RLU in cells-only control wells. Fifty and 80 percent neutralization titers (ND50 and ND80) were estimated using the *nCal* and *drc* packages in R. RLU was first transformed to neutralization using the formula neut = 1 – ([RLU_sample_ – bkgd] / [RLU_VO_ – bkgd]). The neutralization vs. dilution curve was then fit with a 4PL model that was used to estimate the dilution at which there would be 50% or 80% neutralization. For samples with all dilutions having <50% neutralization the result was right censored at the highest concentration. Patient demographic information (sex and age) was extracted from a RedCap survey database. Abbott assay results (including index value) were extracted from the laboratory information system (Sunquest Laboratory).

Correlations were estimated between pairs of neutralization or binding antibody readouts using Pearson’s correlation coefficient (*r*); measures in units of neutralization and IgG concentration were logged prior to estimating correlation. Log-transformed ND50 values and IgG concentrations were approximately normally distributed with few outliers and a low level of censoring, justifying use of Pearson’s correlation and linear regression. Left censored values were given a value of half the level of detection, which corresponded to the first dilution for each neutralization assay. Student’s *t* test was also on log-transformed values. Association of neutralization and IgG concentration with age and BMI were conducted using Spearman’s rank-based correlation and Wilcoxon’s rank-sum tests were used to compare neutralization and IgG in two groups of individuals (e.g. gender, presence of symptom score >2). Statistical significance was determined based on a p-value < 0.05.

## RESULTS

### Cohort characteristics, demographics, survey participation, and serology clinical testing

A total of 1,359 email invitations were sent to 2,655 phase 1 study volunteers and 63 phase 2 volunteers. Among phase 1 and 2 volunteers invited to participate, 973 (72%) and 53 (84%) people consented and completed the enrollment survey. Of these, 967 participants presented for specimen collection between May 4-19, 2020, and 222 (22.8 %) with blood drawn had IgG antibodies to SARS-CoV-2 nucleoprotein according to the Abbott Architect test (index value ≥1.40).Out of these, we randomly selected forty positive samples to evaluate different platforms of SARS-CoV-2 neutralizing antibody assays. Participants had a median age of 51.5 years and a range between 23 and 81 years (Table 1). According to the survey, only one participant reported being hospitalized and four participants (10%) were self-described as asymptomatic. Among participants reporting different symptoms (Table S1), 57.5% had fever while fatigue (87.5%), cough (72.5%), headache (67.5%) and chills (65%) were more prevalent. Based on this, our cohort can be described as representing mild-to-moderate symptomatic infections.

**Table 1.** Demographic and exposure/symptom characteristics of study participants

| **Age** | **N** | **%** |
| --- | --- | --- |
| 23-40 | 8 | 20 |
| 41-50 | 11 | 27.5 |
| 51-60 | 11 | 27.5 |
| 61-70 | 6 | 15 |
| >70 | 4 | 10 |
| Median | 51.5 |  |
| Range | 23-81 |  |
| **Gender** |  |  |
| Female | 16 | 40 |
| Male | 24 | 60 |
| **Exposures/symptoms** |  |  |
| Tested positive | 8 | 20 |
| Symptomatic contact of known positive | 9 | 22.5 |
| Symptomatic without confirmation | 19 | 47.5 |
| Asymptomatic contact of someone symptomatic | 2 | 5 |
| Asymptomatic, no exposures | 2 | 5 |
| Travel outside US since 12/1 | 7 | 17.5 |
| **Other** |  |  |
| Essential worker | 6 | 15 |
| Lives with children | 14 | 35 |

### Cell-based assays provided comparable estimates of neutralization activity

We tested forty heat-inactivated plasma samples in four different cell-based neutralization assays each with a different pair of target cells and virus (Table 2): live recombinant SARS-CoV-2 (rSARS-CoV-2-nLuc) with Vero E6 cells, a lentivirus pseudotyped by SARS-CoV-2 spike (LV-pseudovirus) with HEK 293T cells expressing human ACE2 (293T/ACE2) or TZM-bl cells expressing both ACE2 and TMPRSS2 (TZM-bl/ACE2/TMPRSS2) and a vesicular stomatitis virus (VSV) pseudotyped by SARS-CoV-2 spike (PsVSV-Luc-D19) with Vero cells; a fifth binding neutralization assay using competitive ELISA to assess inhibition of binding of hACE2 and SARS-CoV-2 RBD.

**Table 2.** SARS-CoV-2 neutralization assay platforms used in the study

| Assay | SARS-CoV-2/VeroE6 | VSV-pseudo/Vero | LV-pseudo/293T | LV-pseudo/TZM-bl | Surrogate Virus Neutralization Test (sVNT) |
| --- | --- | --- | --- | --- | --- |
| Cell line | Vero E6 | Vero | HEK293T | TZM-bl | None |
| ACE2 expression | Endogenous | Endogenous | Engineered | Engineered | Recombinant |
| TMPRSS2 expression | No | No | No | Engineered | N/A |
| Virus used for assay | rSARS-CoV-2-nLuc | PsVSV-Luc-D19 | LV-pseudo | LV-pseudo | N/A |
| Virus type | live recombinant | VSV(G*ΔG-luciferase) pseudotyped | pCMV-ΔR8.2 lentiviral packaging with pHR'-CMV-Luc | pSG3ΔEnv lentiviral packaging | N/A |
| SARS-CoV-2 strain/isolate | WA-CDC-WA1-A12/2020 | Wuhan-Hu-1 | Wuhan-Hu-1 (VRC7480) | Wuhan-Hu-1 (VRC7480) D614G | Unknown |
| GenBank | MT020880.1 | MN908947.3 | MN908947.3 | MN908947.3 | N/A |
| position 614 | D | D | G | G | Unknown |

Fifty percent neutralizing dilution (ND50) is a standard numerical parameter to compare virus-neutralizing potency between different samples and studies. To reflect the ultimate capacity of serum antibodies to neutralize virus both ND50 and ND80, the dilutions at which 50% and 80% neutralization is observed, are used together. We serially diluted plasma samples to generate titration curves and estimated ND50 and ND80 relative to positive and negative controls (Fig. S1–S3). All four cell-based neutralization assays performed comparably and generated titration curves necessary for ND50 and ND80 estimation using a four-parameter logistic model (Fig. S1–S3). On average, the slope parameter for neutralization curves with rSARS-CoV-2-nLuc was higher compared to other assays (slope B=3.3 vs. 0.6, 1.4, 1.5 for LV-pseudo/293T, LV-pseudo/TZM-bl and VSV-pseudo/Vero, respectively; all p < 0.001). Geometric mean ND50 from the assay with rSARS-CoV-2-nLuc (141, 95% CI 94-213) did not differ (p=0.2) from the LV-pseudo/293T assay (178, 95%CI 112-283; Fig. 1A, Fig. S4A). However, the LV-pseudo/TZM-bl assay showed significantly lower geometric mean ND50s compared to LV-pseudo/293T (Fig. 1A, Fig. S4A). The VSV-pseudo/Vero assay produced significantly higher ND50 values (geometric mean ND50 of 310, 95%CI 211-454) compared to both the SARS-CoV-2/VeroE6 assay and the two LV-pseudovirus assays (Fig. S4A) suggesting that it is easier to neutralize VSV-based pseudovirus in Vero cells compared to other approaches.

**Figure 1.**
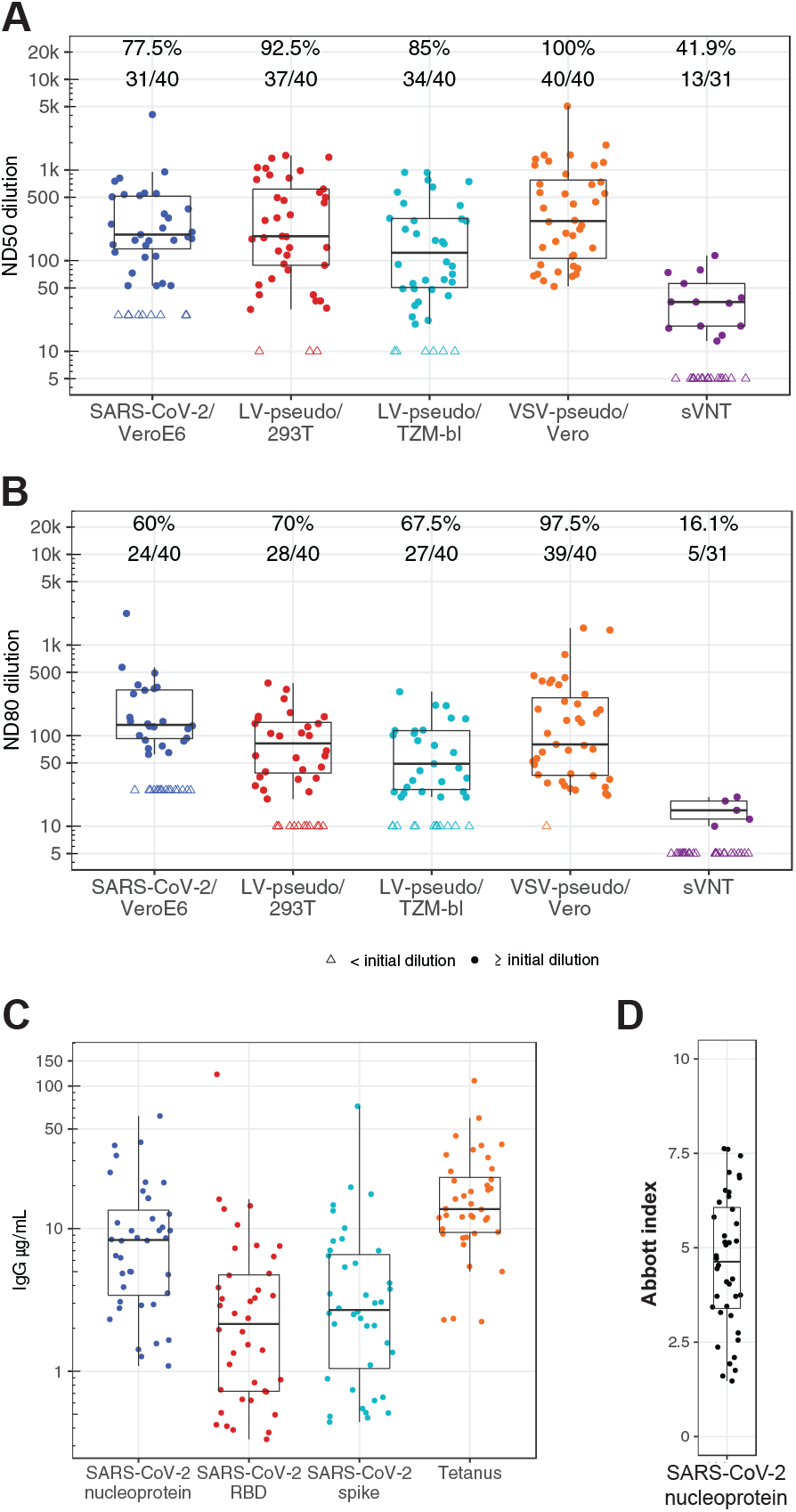
Boxplots of SARS-CoV-2 neutralization and binding antibody concentration for 40 plasma samples from 40 COVID-19 convalescent patients. (A) ND50 and (B) ND80 neutralization titer measured using five SARS-CoV-2 neutralization assays. Each assay defined its own lower limit of detect (LOD) based on the initial dilution: 50-fold for SARS-CoV-2/VeroE6, 20 for the LV and VSV pseudovirus assays and 10 for the sVNT. Data below the LOD (open triangle) is plotted at LOD/2. Number and percent of samples above the LOD is indicated above each plot. (C) Antigen-specific IgG concentration measured using a Luminex bead-based assay. (D) Index values for each sample from the Abbott Architect nucleoprotein IgG assay. For each assay the box represents the extend of the inter-quartile range (IQR) with a line indicating the median; whiskers extend to 1.5 times the IQR.

The assay platforms also differed in their capacity to detect neutralization. In the live-virus assay, LV-pseudo/293T and LV-pseudo/TZM-bl neutralization was detectable at the lowest dilution for 31, 37, and 34 samples, respectively, and therefore permitted estimation of the ND50; ND50 of the remaining samples was censored at the lowest dilution (Fig. 1A). In contrast, VSV-pseudo/Vero permitted estimation of ND50 for all 40 samples (Fig. 1A). With the sVNT, only 31 of 40 samples showed neutralization above 20%, a negative cutoff value according to the manufacturer’s protocol, at the lowest dilution 1:10 (Fig. S3A). To estimate ND50, we selected 13 of these 31 samples and tested them in serial dilutions (Fig. 1A, Fig. S3B). Samples were selected to represent different percent neutralization observed at 1:10 dilution. The resulting ND50 was significantly lower (29.5 95%CI 18.2-47.9) than in the cell-based assays, further supporting the conclusion that the surrogate assay had lower sensitivity compared to the cell-based assays.

We used the same 4PL models to estimate ND80 titers. Though ND80 was consistently lower, the correlation with ND50 was high ranging from Pearson’s r=0.87 for the live-virus assay to r=0.97 for the VSV-pseudovirus assay. Similar to ND50, the ND80 titers also differed among the assays (Fig. 1B, Fig. S4B) with the SARS-CoV-2/VeroE6 assay reporting the lowest number of samples with ND80 titers above the limit of detection (24/40) and VSV-pseudovirus assay showing the highest (39/40). The live-virus assay also showed the smallest difference between ND50 and ND80 titers (Fig. S5, Table S2), a direct consequence of the steeper titration curves observed for this assay (Fig. S1A). For pseudovirus-utilizing assays the difference between ND50 and ND80 was greater and ranged between 2.7 and 4.2-fold. Due to inability to reach 80% neutralization for the many samples in sVNT, we could not calculate ND80 (Fig. S3).

### Strong correlation among neutralization assays

Next, we conducted a correlation analysis of the ND50 and ND80 values derived from each of the five neutralization assays (Fig. 2, Fig. S6). The live-virus and all three pseudovirus neutralization assays generated ND50 values that were highly correlated across samples (Pearson *r* = 0.81 – 0.89) with the highest correlation observed between the two LV-pseudovirus assays (Pearson *r* = 0.89, 95% CI [0.81, 0.94], p<0.001). ND50 reported by the sVNT showed the lowest correlation with the ND50 values from the cell-based assays (Pearson *r* = 0.41 – 0.60). In contrast, correlation was greater between the percent neutralization measured at 1:10 dilution and the outcomes of the cell-based assays (Pearson *r* = 0.73 – 0.80); correlation between ND50 and percent neutralization at 1:10 dilution was modest (*r* = 0.59).

**Figure 2.**
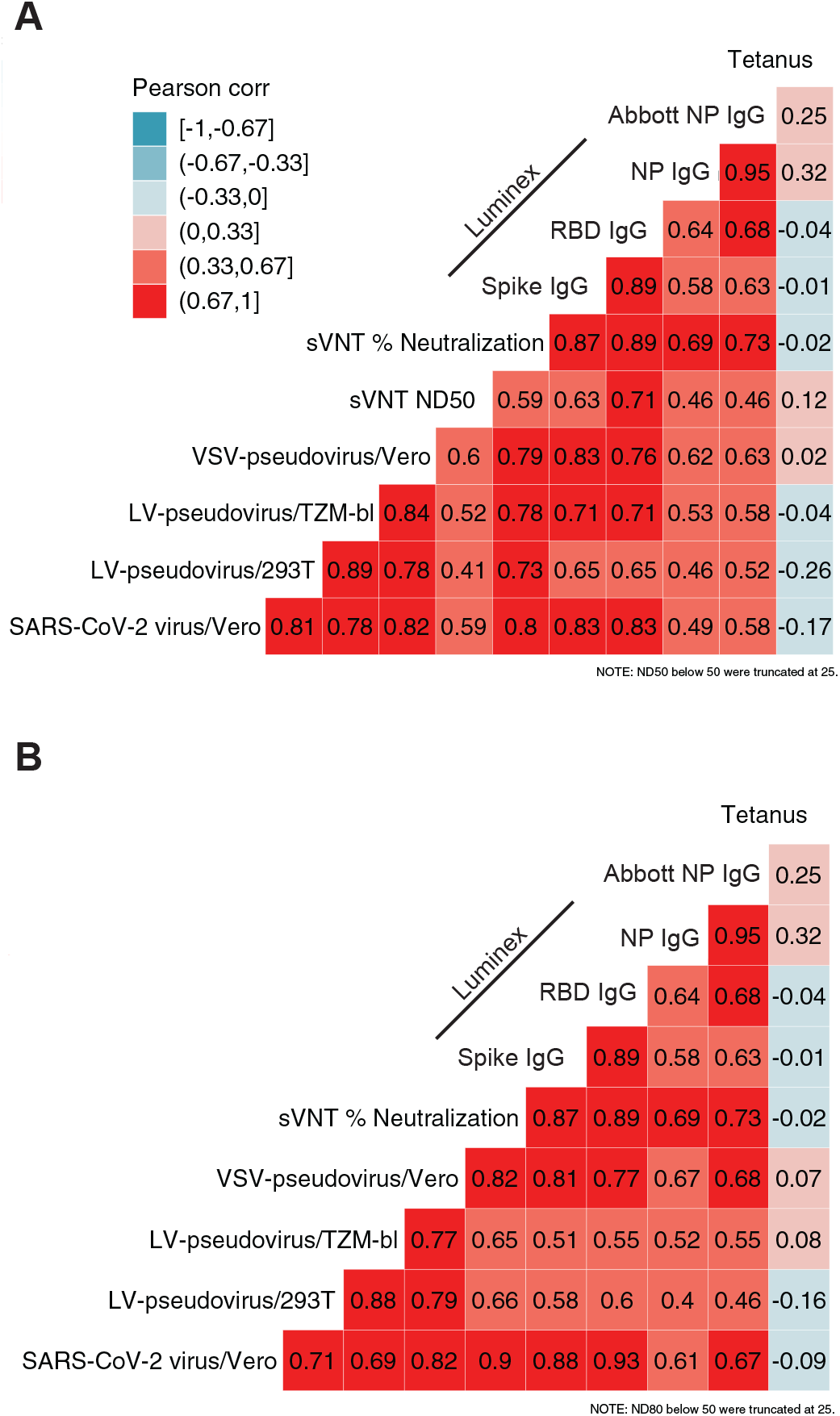
Correlation among assay readouts measuring neutralization or antigen-specific IgG concentration in plasma. Heatmap color is determined by the Pearson’s correlation coefficient (*r*, annotations). Each panel includes either ND50 titers (A) or ND80 titers (B) and their correlation with sVNT % neutralization, SARS-CoV-2 specific IgG concentration (Luminex bead-based assay), the quantitative index of the Abbott nucleoprotein assay and tetanus toxoid-specific IgG concentration.

Similar correlation was observed for ND80 outcomes for cell-based assays (Fig. 2B) with Person’s *r* ranging between 0.69 – 0.88. Only a few samples showed neutralization greater than 80% in sVNT (Fig. S3) and thus correlation between sVNT ND80 and other assays was not estimated.

### Plasma concentration of IgG binding antibodies correlated with neutralization potency

Prior studies of SARS-CoV-2 individuals showed that the serum titer of spike and RBD-binding IgG antibodies was a correlate of neutralizing potency (14, 34, 35). Using quantitative, Luminex-based immunoassay, we measured concentration of IgG to SARS-CoV-2 spike, RBD and nucleoprotein in each of the serum samples; IgG to tetanus toxoid was also measured as a proxy for overall IgG level and state of humoral immunity (Fig. 1C). The mean concentration of nucleoprotein-specific IgG measured in the Luminex assay was 7.3 µg/ml (95%CI [5.3, 10]) while the concentration of both spike and RBD IgG was lower at 2.8 (95%CI [1.9 – 4.1]) and 2.1 µg/ml (95%CI [1.4, 3.3]), respectively. Concentration of tetanus-specific IgG was higher than IgG to SARS-CoV-2 antigens for all individuals (mean 14.5, 95%CI [11.1, 18.9]). Although Abbott SARS-CoV-2 IgG assay is designed and used for qualitative detection of IgG against the SARS-CoV-2 nucleoprotein, the instrument reports index values that can be used in quantitative analyses (Fig. 1D).

Pearson correlation analysis revealed that levels of RBD and spike IgG correlated strongly (Pearson’s *r* = 0.89, 95% CI [0.81, 0.94]) (Fig. 2A). Nucleoprotein-specific IgG measured in our Luminex immunoassay was highly correlated with the quantitative index of the Abbott Architect nucleoprotein IgG assay (Pearson’s *r* = 0.95, 95% CI [0.91, 0.97]). Correlation between levels of nucleoprotein IgG measured either in Luminex or Abbott assay moderately correlated with concentration of spike and RBD-specific IgG with Abbott indexes showing higher *r* values (Pearson’s *r* ranging 0.58 –0.68). There was no significant correlation of tetanus-specific IgG with SARS-CoV-2 spike, spike RBD or nucleoprotein IgG (all p > 0.05).

We then examined the relationship between concentration of SARS-CoV-2 IgG and virus neutralization (Fig. 2, Fig. S7). We found that IgG concentrations to each of the SARS-CoV-2 antigen was positively correlated with neutralization potency measured with each of the neutralization assays (Pearson’s *r* = 0.46 – 0.83; Fig. 2A). The correlation of live-virus neutralization ND50 with concentration of spike (Pearson *r* = 0.83, 95% CI [0.7, 0.91]) and RBD-specific (0.83 95% CI [0.7, 0.91]) IgG was comparable to the correlations observed with the other neutralization assays. Nucleoprotein-specific IgG concentration as well as the quantitative index from the Abbott test were only moderately correlated with ND50 obtained in cell-based assays, with *r* ranging between 0.46 – 0.63. In contrast the correlation of nucleoprotein-specific IgG was higher with the percent neutralization measured with the sVNT (Luminex, Pearson’s *r* = 0.69, 95% CI [0.49, 0.83] and Abbott, *r* = 0.76, 95% CI [0.54, 0.85]). Tetanus-specific IgG did not correlate with any of SARS-CoV-2-associated IgG concentrations or neutralization titers.

### Effect of age, gender and disease symptoms

We explored if demographic and physiologic parameters were associated with SARS-CoV-2 specific IgG or neutralization. Previously, body mass index (BMI), female sex and age were reported to positively correlate with antibody titers against SARS-CoV-2 (36). We asked whether ND50 titers obtained from each neutralization assay correlated with age, gender, BMI or self-reported disease symptoms (Table 3). We found a moderate positive correlation between age and concentrations of spike-specific (Spearman’s rho=0.37, p=0.02), spike RBD-specific (rho=0.39, p=0.013) and nucleoprotein-specific (rho=0.45, p=0.003) IgG. Similarly, there were positive correlations between age and neutralization titer, though the correlations tended to be higher with ND80 compared to ND50 titer (Fig. 3). For example, the correlation coefficient of age with live-virus neutralization ND80 was rho=0.51 (p=0.001), compared to rho=0.28 (p=0.075) for ND50. No consistent significant correlations with BMI, gender or symptoms were observed (Table 3).

**Table 3.** Tests for association of SARS-CoV-2 antibody neutralization and binding with age, BMI, sex and symptoms

| Assay | Measure | N | BMI<br>rho<sup>1</sup> (p-value) | Age<br>rho<sup>1</sup> (p-value) | Sex (female)<br>fold-difference<sup>2</sup> (p-value) | No. of symptoms<br>(of 19 surveyed)<br>rho<sup>1</sup> (p-value) |
| --- | --- | --- | --- | --- | --- | --- |
| SARS-CoV-2/VeroE6 | ND50 | 40 | 0.20 (0.2107) | 0.28 (0.0751) | 0.52 (0.1646) | 0.08 (0.6248) |
| VSV-pseudo/Vero | ND50 | 40 | 0.33 (0.0376) | 0.30 (0.0602) | 0.44 (0.0317) | -0.13 (0.4225) |
| LV-pseudo/293T | ND50 | 40 | 0.22 (0.1713) | 0.24 (0.1288) | 0.52 (0.1467) | 0.04 (0.8043) |
| LV-pseudo/TZM-bl | ND50 | 40 | 0.38 (0.0164) | 0.27 (0.0885) | 0.57 (0.2239) | 0.10 (0.5395) |
| sVNT neutralization | ND50 | 31 | 0.40 (0.0252) | 0.43 (0.0160) | 0.64 (0.2543) | 0.16 (0.3896) |
| SARS-CoV-2/VeroE6 | ND80 | 40 | 0.31 (0.0500) | 0.51 (0.0007) | 0.64 (0.2362) | 0.04 (0.8239) |
| VSV-pseudo/Vero | ND80 | 40 | 0.37 (0.0181) | 0.32 (0.0466) | 0.40 (0.0360) | -0.11 (0.4967) |
| LV-pseudo/293T | ND80 | 40 | 0.36 (0.0212) | 0.32 (0.0444) | 0.54 (0.1411) | 0.07 (0.6798) |
| LV-pseudo/TZM-bl | ND80 | 40 | 0.30 (0.0612) | 0.29 (0.0738) | 0.65 (0.3360) | 0.10 (0.5347) |
| sVNT neutralization | ND80 | 31 | 0.09 (0.6146) | 0.50 (0.0038) | 0.81 (0.1806) | -0.17 (0.3675) |
| sVNT neutralization (1:10 dilution) | % | 40 | 0.33 (0.0374) | 0.40 (0.0106) | 0.87 (0.3075) | 0.09 (0.5670) |
| Abbott nucleoprotein | index | 40 | 0.29 (0.0719) | 0.45 (0.0034) | 0.71 (0.0318) | -0.04 (0.8021) |
| SARS-CoV-2 spike-specific IgG | µg/mL | 40 | 0.30 (0.0605) | 0.37 (0.0197) | 0.64 (0.2790) | 0.09 (0.5779) |
| SARS-CoV-2 RBD-specific IgG | µg/mL | 40 | 0.28 (0.0849) | 0.45 (0.0035) | 0.69 (0.4522) | 0.16 (0.3359) |
| SARS-CoV-2 nucleoprotein-specific IgG | µg/mL | 40 | 0.28 (0.0830) | 0.39 (0.0126) | 0.47 (0.0415) | -0.13 (0.4096) |
| Tetanus toxoid-specific IgG | µg/mL | 40 | 0.10 (0.5381) | -0.14 (0.3853) | 0.85 (0.8166) | -0.12 (0.4576) |

<sup>1</sup>Spearman's rank correlation coefficient
<sup>2</sup>fold-difference indicates the geometric mean value in females/males
<sup>3</sup>fold-difference indicates the geometric mean value in volunteers/males

**Figure 3.**
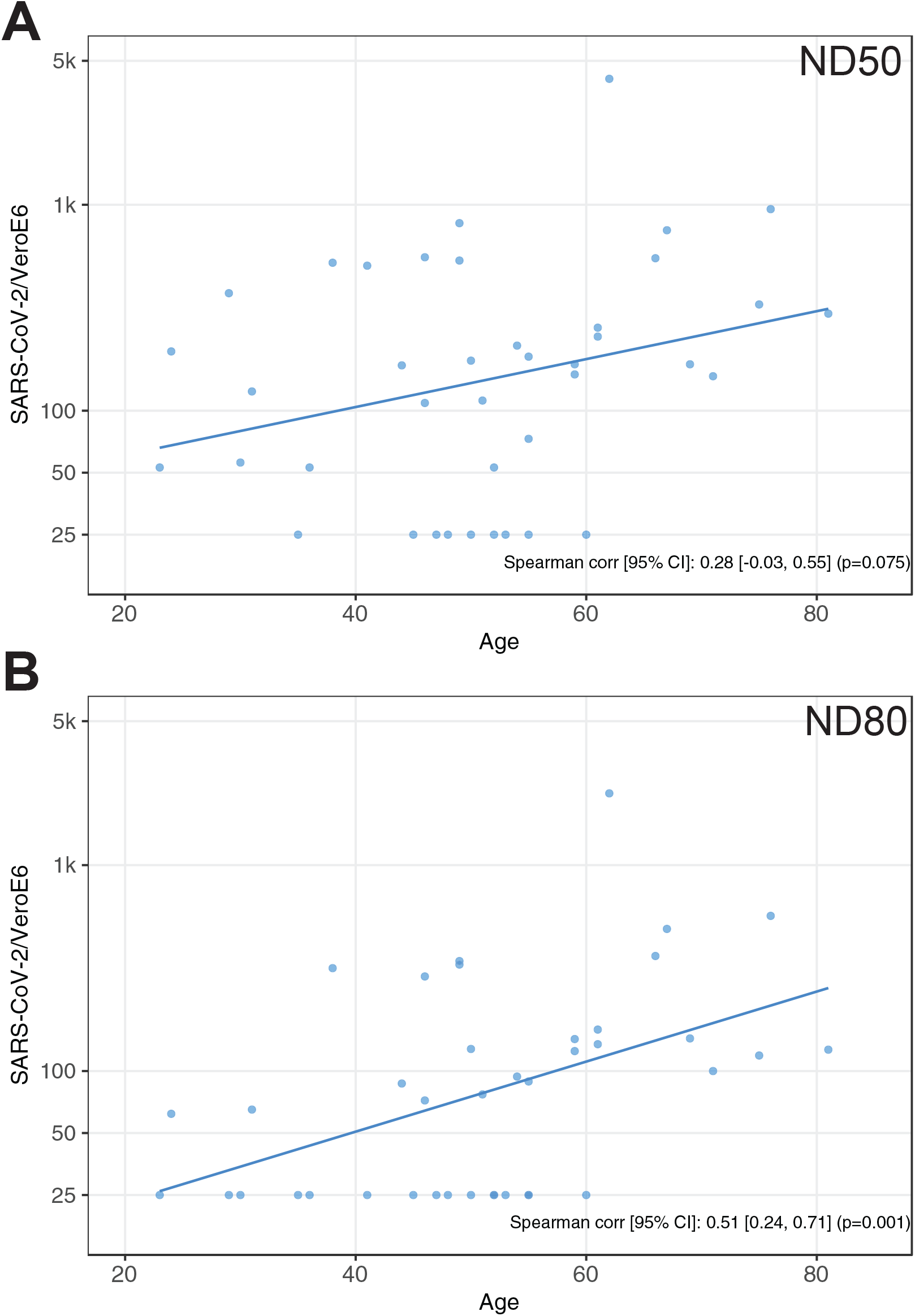
Correlation analysis of plasma neutralizing potency and age of participants. A, ND50 versus age; B, ND80 versus age.

## Discussion

In this study we conducted a detailed comparison of different SARS-CoV-2 neutralization assays using a set of 40 plasma samples collected from SARS-CoV-2 convalescent individuals with mild-to-moderate disease involved in a county-wide outbreak of COVID-19. These data show a high level of congruency among cell-based SARS-CoV-2 neutralization assays. The 50% and 80% neutralization titer readouts of cell-based assays were highly correlated with each other and with the concentration of RBD and spike-specific IgG. The results of the ELISA-based sVNT were also positive correlated with the other neutralization assays, however the correlation was modest in comparison. Though levels of spike-specific IgG were highly correlated with neutralization, this does not indicate that all spike-specific binding IgG have neutralization activity, rather it implies that individuals who produce spike-specific binding antibodies are also likely to make neutralizing IgG. The correlation between nucleoprotein-specific IgG and neutralization was consistently lower than the correlations of spike and spike RBD-specific IgG with neutralization. This is consistent with known mechanisms of neutralization, which involve binding and/or blocking the spike:ACE2 receptor binding domain; the moderate correlations of nucleoprotein-specific IgG with neutralization may indicate that presence of any SARS-CoV-2 specific IgG is a biomarker of the presence of neutralizing antibodies as well. The association of age with both the plasma concentration of spike-specific IgG and neutralization titer suggests that the previously reported association of high neutralization titer among older individuals may be mediated by higher concentrations of spike and spike RBD-specific IgG. Whether this is a result of prior infections with seasonal coronaviruses or an effect of age on the developing immune response to SARS-CoV-2 is not clear yet.

Previously, in a cohort of severely ill COVID-19 patients, deceased individuals were reported to have higher concentrations of nucleoprotein-than spike- and RBD-specific IgG, and the opposite scenario was associated with survival (37). In our study, individuals with mild-to-moderate disease also demonstrated higher concentrations of nucleoprotein IgG compared to spike and RBD IgG suggesting that the immune response to spike and nucleoprotein differ in milder forms of disease. Of interest, we did not see any correlation between the neutralizing potency of plasma and BMI. As our cohort was largely uniform with regard to disease severity, our study cannot comment on the association between spike and nucleoprotein antibodies and disease severity.

With the set of cell lines used for the assays in this study, we were able to address questions regarding the influence of proteolytic cleavage of SARS-CoV-2 spike on virus neutralization by serum antibodies which is important for choosing an assay that would provide more physiologically relevant outcomes. Although there was no significant difference observed in virus titer at 48 h post infection on the wildtype Vero cells and Vero cells expressing furin, at the early time point cells expressing protease showed a higher virus titer (9). Proteolytic cleavage was also shown to be essential for SARS-CoV-2 infectivity on other cell types (9, 17). Therefore, cell lines expressing TMPRSS2 or a related protease would allow testing for the possible role of proteolytic cleavage of the spike glycoprotein in virus infectivity. In contrast to this reasoning, comparison of different cell types used for pseudovirus assays revealed that the presence of TMPRSS2 is not critical for assay performance. As such, TZM-bl cells designed to express both ACE2 and TMPRSS2 showed no significant difference compared to 293T cells that do not express TMPRSS2 endogenously and were only expressing ACE2. TZM-bl cells are widely used in HIV research for neutralization assays with both live viruses and pseudoviruses due to the assay robustness and reproducibility (38, 39).

A neutralization platform based on Vero cells and VSV-pseudovirus demonstrated the highest sensitivity among assays tested. This could be explained by lower affinity of interaction between SARS-CoV-2 and simian ACE compared to the human ACE2 and by different density of spike glycoprotein on the surface of VSV pseudovirus. While increased sensitivity may lead to overestimation of neutralization potency, it can also be useful for specimens with low neutralizing activity or when sample volume is limited such as for mucosal secretions and washes. However, the high correlation between the live-virus assay and Vero/VSV-pseudovirus assay suggests that data obtained in the latter can accurately reflect the sample potency to neutralize wildtype SARS-CoV-2.

ELISA-based assays have two major limitations: i) inability to account for synergistic action of antibodies targeting different epitopes; and ii) detection only of antibodies that block interaction between RBD and ACE2, thus omitting antibodies that neutralize virus via non-RBD sites on the virus glycoprotein (24, 40). For example, synergistic action of antibodies against RBD and the S2 domain has been reported (41). There are two ways of performing such a surrogate assay: soluble biotinylated ACE2 competing with serum antibodies for binding to immobilized RBD or spike, or an opposite version with soluble RBD and immobilized ACE2 (21). Abe et al. found that an assay with soluble ACE2 and immobilized RBD was more sensitive and yielded ND50 values that correlated with ND50 titers obtained in the classical cell-based PRNT with a coefficient of determination of 0.6. The GenScript assay that we have tested in our study utilizes immobilized ACE2, which likely explains why we were not able to measure ND50 titers for the majority of samples. Of note, samples used by Abe et al. were also collected from mild-to-moderate COVID-19 patients. In conclusion, a surrogate assay can be used cautiously as an alternative to cell-based assays to obtain preliminary qualitative results, to rapidly distinguish between samples with high and low neutralizing potency, and when a cell-based assay is not available or reasonably feasible.

## Data Availability

N/A

## Acknowledgements

We thank Dr. Mindy Minor for critical reading of the manuscript and editorial help, X for technical assistance, and Sara Thiebaud for assistance with data management and analysis. This work was funded by: NIAID Service Agreement 225472-99 to RKS, R01AI134878 and UM1AI068614 to LC, Fred Hutch Evergreen grant to AMS.

