## Supplemental material for "Evaluation of SARS-CoV-2 neutralization assays for antibody monitoring in natural infection and vaccine trials"

**Supplementary materials**

Supplementary Table 1. Symptoms reported by study participants

| Symptom | Yes | No | Missing | Percent reporting symptom |
|---|---|---|---|---|
| **fever** | 23 | 11 | 6 | 57.5 |
| **chills** | 26 | 10 | 4 | 65 |
| **fatigue** | 35 | 4 | 1 | 87.5 |
| **myalgia** | 27 | 9 | 4 | 67.5 |
| **sore throat** | 20 | 14 | 6 | 50 |
| **cough** | 29 | 6 | 5 | 72.5 |
| **rhinorrhea** | 24 | 10 | 6 | 60 |
| **dyspnea** | 22 | 13 | 5 | 55 |
| **wheezing** | 6 | 19 | 15 | 15 |
| **chest pain** | 13 | 16 | 11 | 32.5 |
| **other respiratory** | 8 | 17 | 15 | 20 |
| **headache** | 27 | 8 | 5 | 67.5 |
| **nausea** | 9 | 18 | 13 | 22.5 |
| **abdominal pain** | 6 | 20 | 14 | 15 |
| **diarrhea** | 14 | 16 | 10 | 35 |
| **loss senses** | 26 | 8 | 6 | 65 |
| **eye pain** | 7 | 19 | 14 | 17.5 |
| **rash feet** | 2 | 21 | 17 | 5 |
| **rash body** | 4 | 21 | 15 | 10 |

**A**

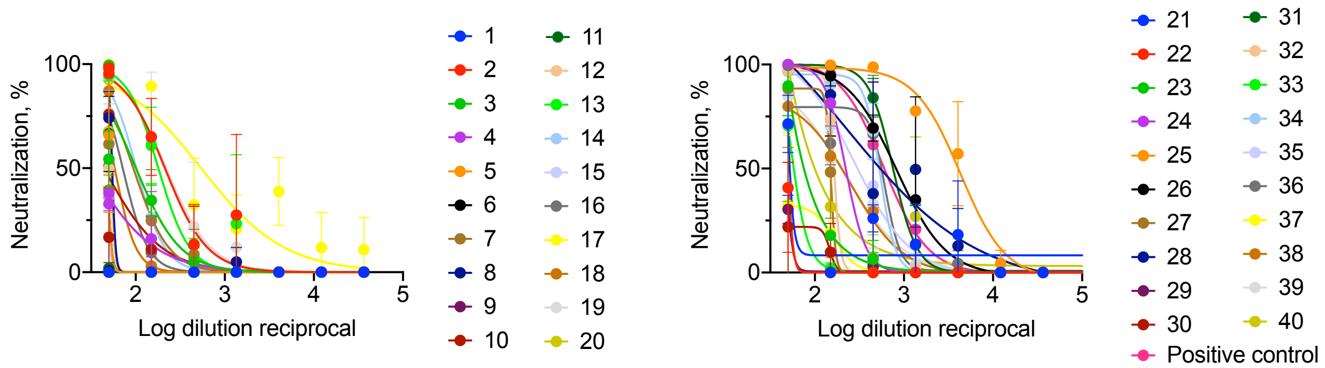


**B**

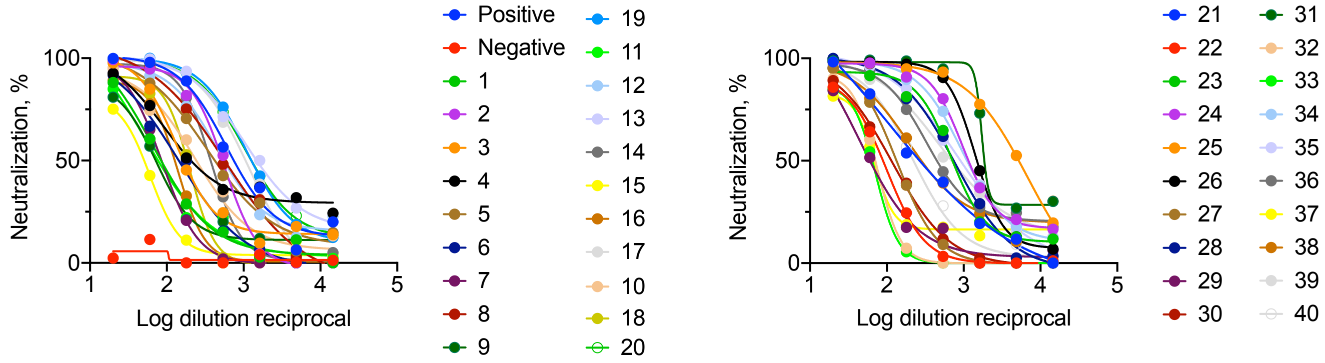


Supplementary Figure 1. Neutralizing antibody assay with SARS-CoV-2-nLuc in Vero E6 cells (A) and with PsVSV-Luc-D19 in Vero cells (B).

**A**

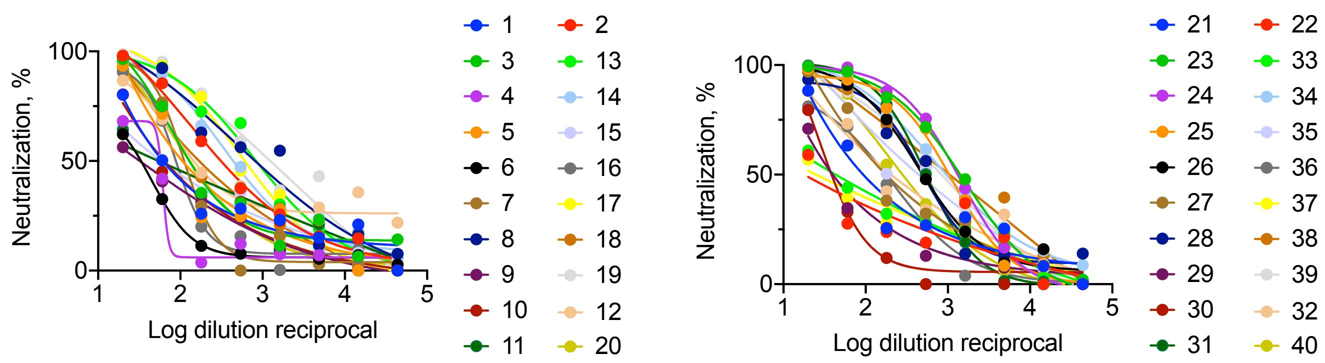


**B**

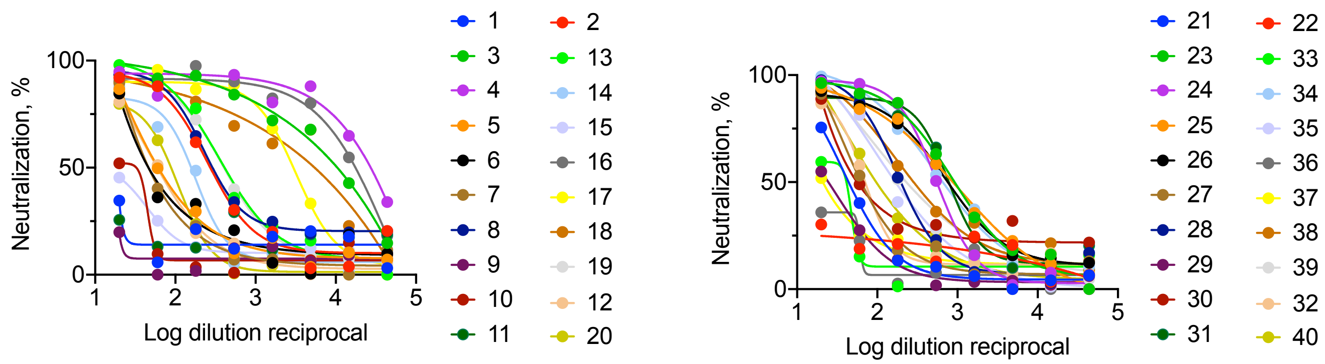


Supplementary Figure 2. Neutralizing antibody assay with LV-pseudovirus in 293T/ACE2 cells (A) and with LV-pseudovirus in TZM-bl/ACE2/TMPRSS2 cells (B).

**A**

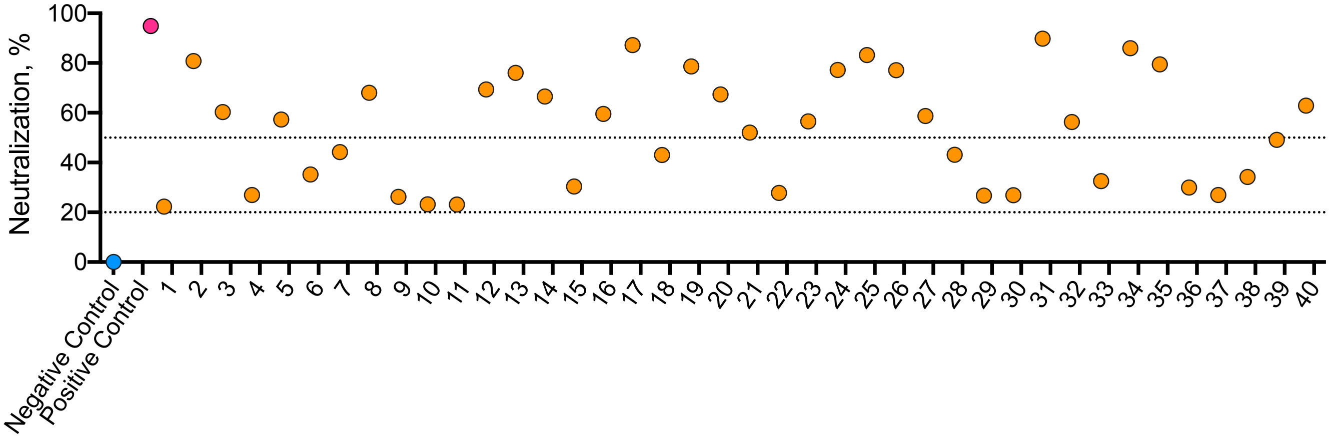


**B**

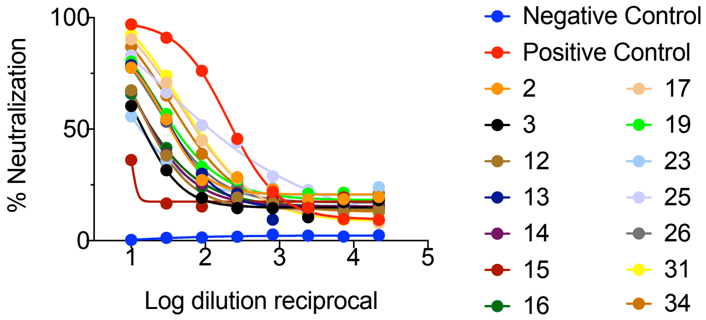


Supplementary Figure 3. Surrogate virus neutralization test from GenScript. A, samples tested in 1:10 dilution as per manufacturer protocol. Dotted lines show 50 and 20% neutralization, respectively. Twenty percent is suggested as positivity cutoff by the manufacturer; B, plasma samples were titrated 2-fold starting at 1:10 to impute ND50 titers.

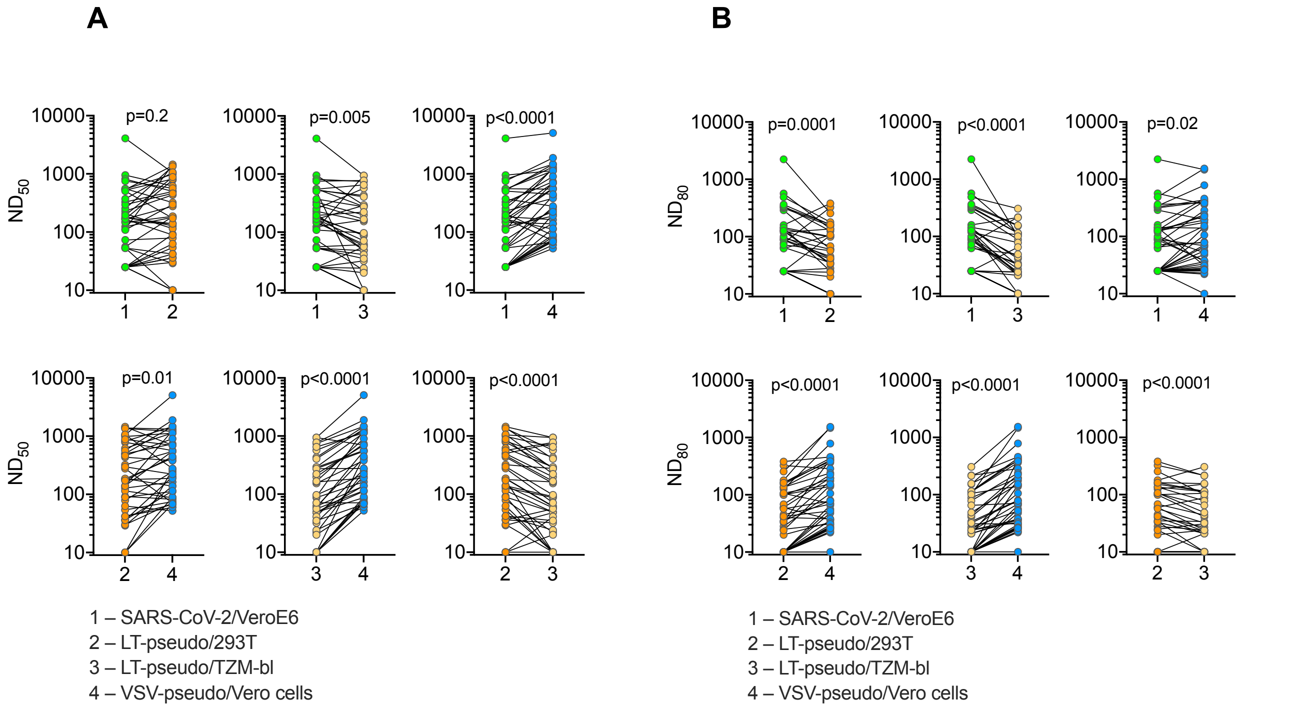


Supplementary Figure 4. Comparison of ND50 (A) and ND80 (B) titers measured in cell-based assays

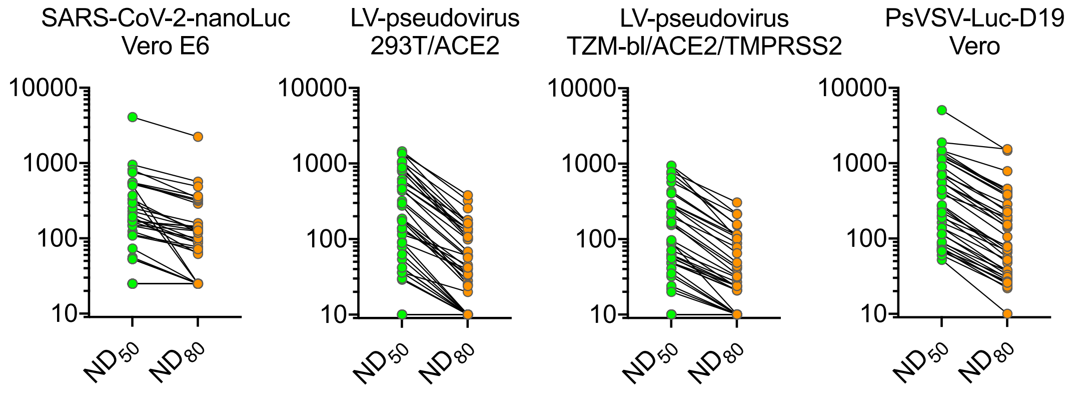


| Assay | $ND_{50}$ | $ND_{80}$ |
|---|---|---|
| SARS-CoV-2-nanoLuc Vero E6 | 141.3 | 79.29 |
| LV-pseudovirus 293T/ACE2 | 177.9 | 41.96 |
| LV-pseudovirus TZM-bl/ACE2/TMPRSS2 | 89.95 | 33.03 |
| PsVSV-Luc-D19 Vero | 309.7 | 102.8 |

Supplementary Figure 5. Pseudovirus assays but not live virus assay allows reliable ND80 measurement. Geometric mean ND50 and ND80 for the corresponding assay are shown underneath each graph

Supplementary Table 2. Fold change between ND50 and ND80 values

| Neutralization | Mean ND fold change [95% CI] | P-value |
|---|---|---|
| SARS-CoV-2/Vero E6 | 1.954 [1.62, 2.29] | <0.0001 |
| LV-pseudovirus/293T | 4.573 [3.65, 5.5] | <0.0001 |
| LV-pseudovirus/TZM-bl | 4.478 [3.48, 5.48] | <0.0001 |
| PsVSV-Luc-D19/Vero | 2.967 [2.7, 3.24] | <0.0001 |

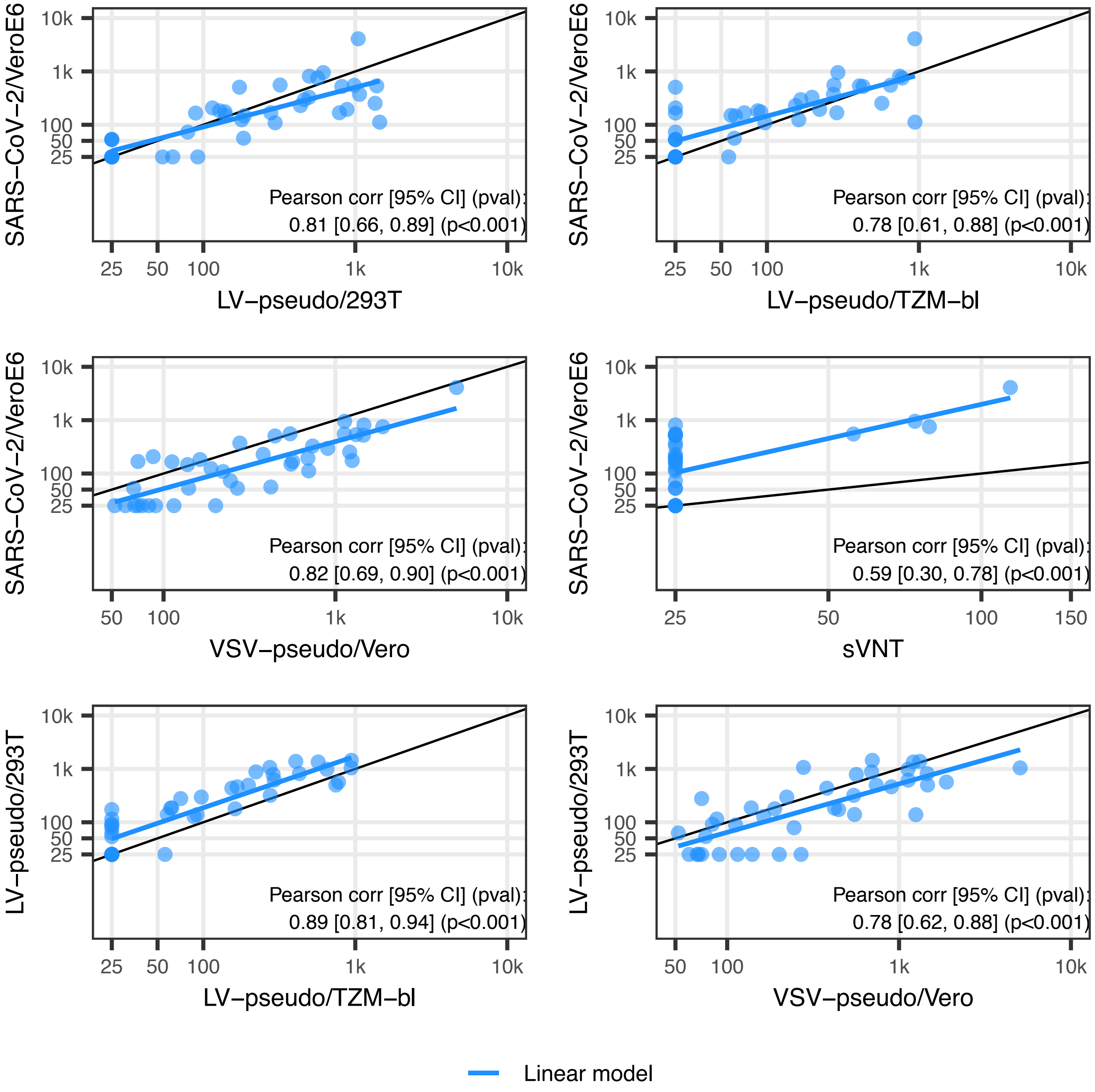


Supplementary Figure 6. Pearson correlation model analysis of ND50 titers among neutralization assays.

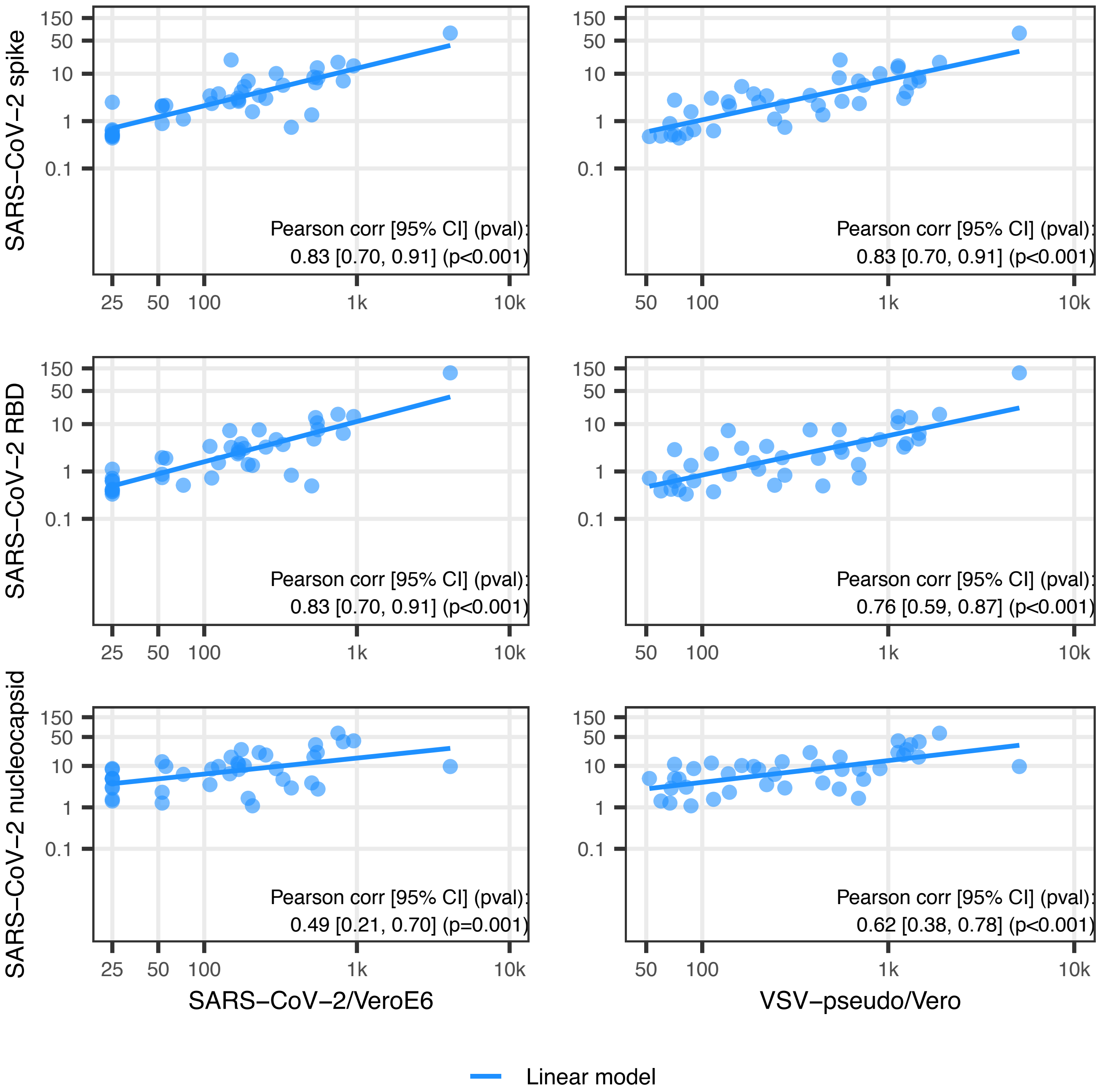


Supplementary Figure 7. Pearson correlation model analysis of ND50 titers vs SARS-CoV-2 specific IgG concentration in plasma samples.
